# A Randomized Controlled Trial Evaluating a Community-Based, Family Network Heart Health Intervention - the SERVE OC Trial: Design, Rationale and Baseline Findings

**DOI:** 10.64898/2026.08.31.26361871

**Authors:** Bernadette Boden-Albala, Jeffrey J. Wing, Matthew J. Landry, Megan Castro, Desiree Gutierrez, Cassandra Cardenas, Julie Rousseau, Amir Rahmani, Aryanna Chavez, Xueting Ding, Alissa Kurzman, Bruce Albala

## Abstract

**Background:** Cardiovascular disease (CVD) disproportionately burdens underserved communities, where social determinants of health (SDOH) perpetuate persistent disparities. Family-based interventions leveraging social support represent a promising yet understudied approach. We describe the rationale, design, and methods of the Skills-based Educational strategies for the Reduction of Vascular Events in Orange County (SERVE OC) RCT and present baseline characteristics of enrolled families.

**Methods:** SERVE OC is a 2-arm RCT of 190 Latino and Vietnamese families (486 individuals) randomized to the family-based intervention or individual self-management. The intervention was grounded in social network theory while employing community engaged strategies. Primary outcomes include achieving ideal cardiovascular health (CVH) defined by AHA Life’s Essential 8 (LE8) and systolic blood pressure reduction at 12, 24, and 36 months. Baseline assessments include demographics, LE8, psychosocial factors, food security, and SDOH. Descriptive statistics and regression analyses examined cohort characteristics and associations between SDOH, food security, and LE8.

**Results:** Over 83% of participants had suboptimal LE8 scores. Average adult total LE8 scores were 66.61 ±11.96, with physical activity as the weakest domain, compared to an average of 76.52±10.15 in children. Greater SDOH burden and food security were associated with significantly lower odds of ideal CVH and lower LE8 scores respectively.

**Conclusions:** SERVE OC demonstrates the feasibility of enrolling families in community-engaged RCT targeting CVD disparities in underserved population. Baseline findings confirm substantial CVD risk and SDOH burden underscoring the need for multi-level, culturally tailored interventions. Trials results will inform scalable, family-focused strategies for CVD prevention across the life course.

**Clinical Trial Registration:** URL: https://www.clinicaltrials.gov/; Unique Identifier: NCT05641519.

**Clinical Perspective:**

- The SERVE OC study evaluates a novel, family-based cardiovascular risk reduction intervention utilizing a community health worker model that strategically leverages social networks to promote sustainable, healthy behavior change.
- The approach fosters support with a focus on optimizing primary and secondary prevention of cardiovascular disease within the family unit.
- Engaging family support systems in clinical prevention strategies may improve overall adherence to lifestyle modifications and risk factor management across diverse communities.

## 1 Introduction

Cardiovascular disease (CVD) is a leading cause of death and disability in the US and globally, and is associated with age, sex, race, ethnic, geographic, and other disparities.^1,2^ In the US, one in three adults receive cardiovascular related care and annual costs have increased from $393 billion to $1490 billion in 2024.^3^ In response, efforts towards increasing CVD prevention and risk reduction have greatly increased, with a critical focus on primary and secondary prevention strategies. However, despite these broader efforts, primary and secondary prevention strategies have yielded few advances.

Optimizing CVD health, including the reduction of disparities, is multifaceted. Interventions need to address biological, social, psychological, and environmental factors, including social determinants of health (SDOH).^4,5^ SDOH examines a community’s social support, neighborhood/environment, education, food and job security, and socioeconomic status and its effects on barriers to access, disparities, risk prevention management, and more.^6^ Significant consideration needs to be given to the mitigation of health disparities faced by vulnerable groups, including immigrants, vulnerable populations, the LGBTQ+ community, the isolated elderly population, and racial-ethnic minority groups.^7,8^ For example, Black, Latino, and Vietnamese communities are faced with greater CVD disparities, including higher prevalence, lack of awareness, poor treatment and control of hypertension.^9–11^ Further, limited physical and socio-economic resources within communities disproportionately hinders community strategies to prevent and manage CVD.^8,10,12,13^ The evidence strongly suggests that implementation of a multi-level approach to mitigating disparities in CVD utilizing methods rooted in achieving health equity to improve health outcomes and general wellbeing for all.^7^ This “roadmap” to reducing CVD disparities emphasizes targets areas to optimize prevention, including partnering with stakeholders through community engagement, and identifying risk factors within the context of SDOH.^7^

Multifaceted approaches to CVD prevention have identified a number of key areas to enhance successful interventions, including community engagement, faith-based organizations, and social networks.^14–16^ Our work has focused on the ways in which social networks may optimize strategies to enhance CVD health. An important and novel component of these network interventions goes beyond targeting risk behaviors in individuals and incorporates social networks as forms of social support.^17,18^ Indeed, there is an extensive body of evidence that links social isolation and a lack of social support to increased morbidity and mortality across various disease specific populations.^19–27^ Strong social support networks have been associated with healthy aging, higher functional status, MI reduction, lower stroke incidence rates, and overall decreased mortality.^19,23–26,28^ Social support may also be directly related to stress-reducing physiological mechanisms.^29–31^ Contrastingly, a lack of functional social support negatively impacts quality of life and physical recovery, while increasing rates of depression.^32^ In the Stroke Warning Information and Faster Treatment (SWIFT) study, social isolation resulted in a longer delay in getting to the emergency room and receiving the necessary treatment.^33,34^ Other large prospective cohort studies demonstrate that social disconnection or isolation significantly elevates post-event vascular risks and overall mortality.^22,35^ Disease interventions that include components of learning, counseling and social support (including support group sessions) may provide layers of reinforcement to an intervention model focused on knowledge retention and behavioral change.^36,37^ Social support intervention strategies that have included the use of patients and family members can promote reinforcement and self-efficacy, which have been exemplified in a number of intervention studies on stroke recovery.^38–41^

Additionally, social support is likely to be even more important, particularly in lower socioeconomic and minority communities where access to health and social services may be limited. Risk factors like hypertension, obesity and high cholesterol may be shared among cohabitating family members or even within wider kinship groups and social support groups.^8,42^ As a result, one family member’s cardiovascular event or risk profile can be leveraged to motivate and galvanize a larger social circle toward making powerful behavioral changes that will improve each member’s risk factor profile, possibly preventing primary cardiovascular events in the process. Leveraging family dynamics, shared environment, and intergenerational support is a multilevel method in family-based interventions that can enhance education and promote healthy lifestyle behavior change.^43,44^ A culturally tailored, family-based intervention for diabetes within the Hispanic community showed a significant decrease in BMI one-month post-intervention, and improvements in systolic blood pressure (SBP) and HbA1c.^44^ The Discharge Educational Strategies for Reduction of Vascular Events (DESERVE) study highlighted the efficacy of utilizing social networks in stroke prevention within underserved communities by reducing SBP by 10mmHg through implementing a skills-based, culturally tailored discharge education intervention.^15^ The DESERVE trial employed a community health worker (CHW) model to deliver culturally relevant presentations and community-initiated narratives that reinforced healthy behavior change and enhanced risk reduction in Latino communities one year post-stroke.^15^ Findings showed stroke patients who had contact with one-to-two or three-to-five friend/family members at least several times a week had a greater reduction in SBP compared to patients who had no weekly contact with any of their friends and family members.^45^ While there are promising results of family-based interventions to mitigate cardiovascular risk factors, there have been few clinical trials examining the role of family social networks in CVD prevention.

Expanding upon DESERVE’s social networks findings to include family-based interventions focused on CVD risk reduction, this study also utilizes community-based participatory research (CBPR) strategies to target disadvantaged populations within an urban setting, our local community of Orange County. This Skills-Based Educational Strategies for Reduction of Vascular Events in Orange County (SERVE OC) clinical trial tested the implementation of a novel family-network intervention compared to individual self-management in elevating Life’s Essential 8 (LE8) scores among family participants from Latino and Vietnamese communities of Santa Ana, Garden Grove, Anaheim, and Westminster, CA.^46^ A second aim tested the intervention in the reduction of blood pressure (BP) in adult family participants. Community engagement strategies involved CHWs that tested the efficacy of a family-based intervention versus usual self-management to address both primary and secondary CVD prevention. Overall, SERVE OC evaluated the efficacy of the family-based SERVE OC intervention versus individual self-management (ISM) in achieving “ideal” cardiovascular health (CVH) using the American Heart Association’s (AHA) LE8 score to evaluate the efficacy of the SERVE OC intervention versus ISM in producing a significant (10 mmHG) decrease in SBP among adults.

## 2 Methods

### 2.1 Study Design and Ethics Statement

SERVE OC was a community-engaged 2 arm randomized controlled trial (RCT) testing the effect of a CHW-led, family-based lifestyle intervention versus self-management in improving the AHA’s LE8 score within Latino and Vietnamese families in Orange County at 12, 24, and 36 months post-enrollment. The culturally tailored intervention strategy utilized CHWs to support families in setting goals, building skills, and implementing risk reduction strategies for hypertension and cardiovascular risk factors. Additionally, all families are given a Withings BPM Connect device, which is a remote BP monitor (RBPM), to collect BP measurements from adult participants at-home throughout the duration of the 3-year study. Ethical approval for this study was granted by the University of California, Irvine Institutional Review Board, and was registered on clinicaltrials.gov (NCT05641519). Figure 1 illustrates a broad overview of the study design, with further details given in the subsequent sections. The data, analytic methods, and study materials that support the findings of this study are available from the corresponding author upon reasonable request.

**Figure 1.**
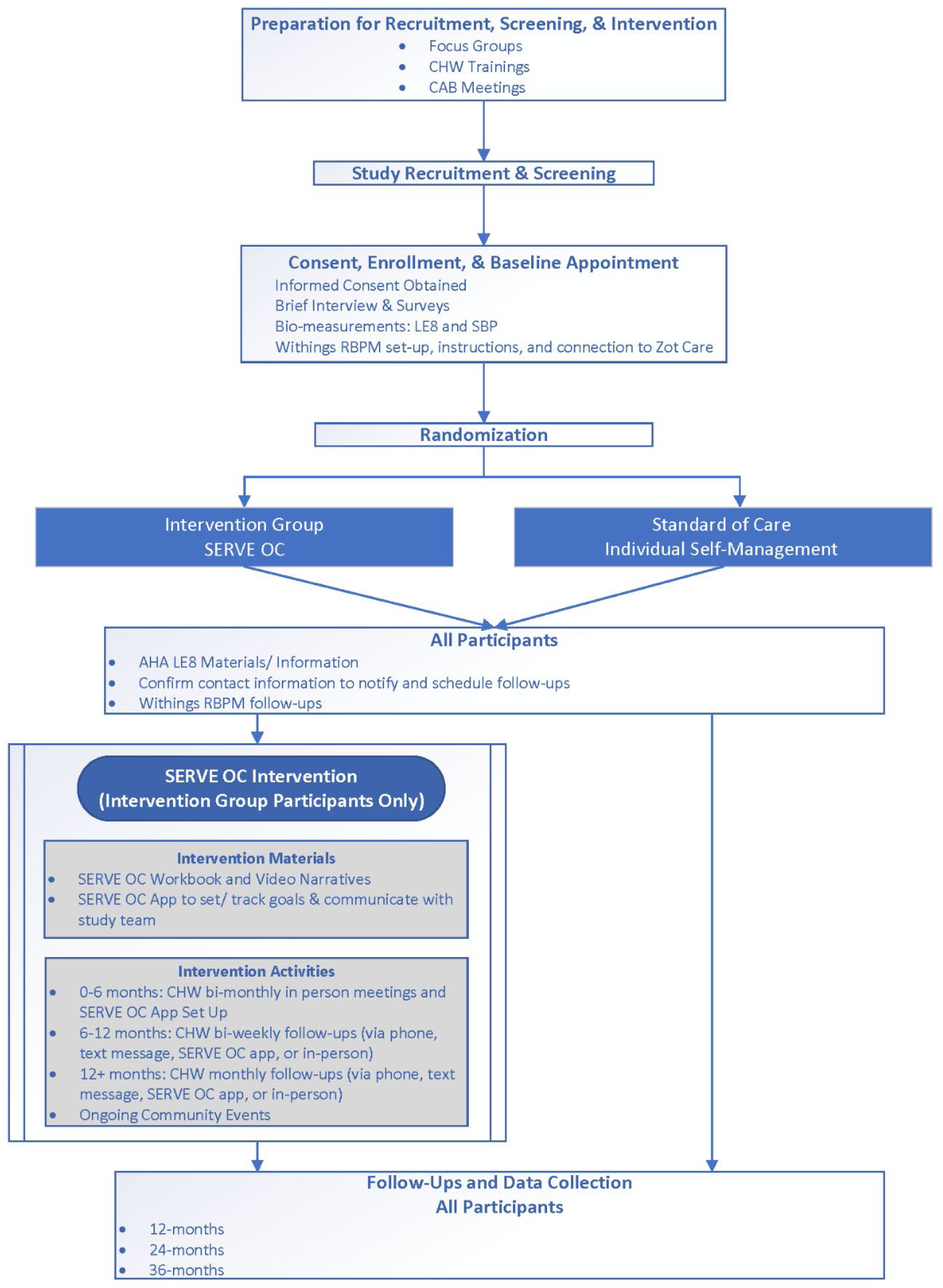
SERVE OC Study Design Flow Chart. Schematic representation of the study phases, including recruitment, eligibility screening, consent and enrollment, randomization baseline data collection, intervention components, and primary outcome assessment points.

#### 2.1.1 Underlying Theoretical Frameworks

The multi-level SERVE OC intervention was based in social networks theory (SNT).^47–49^ SNT provides a crucial lens for examining how social relationships influence health, and for this study – CVH. SNT posits that social support networks profoundly impact health through 4 pathways: companionship, social influence, social support, and social undermining.^47^ The SERVE OC intervention sought to leverage family social network functions to optimize vascular health, which can be visualized in Figure 2.^47^ The SNT is used to map and analyze the structure of family and community networks, identifying key individuals and the flow of social support.^47^ CVH outcomes are influenced by behavioral lifestyle changes, and we tested whether family members who are positively engaged can help facilitate positive reinforcement of these behavioral changes throughout the network.^30,31,45^ Because of the multidimensional nature of the intervention, SERVE OC integrated the socioecological model (SEM) to extend its impact to community and policy levels, aiming for broader systemic change.^49^ The SEM framework is used to identify community and policy-level barriers, including access to care and other factors of SDOH, which would tailor the intervention to the specific needs of Latino and Vietnamese communities. These two theories work together to create a multilevel intervention focused on improving CVH outcomes at the individual, family, and community level.

**Figure 2.**
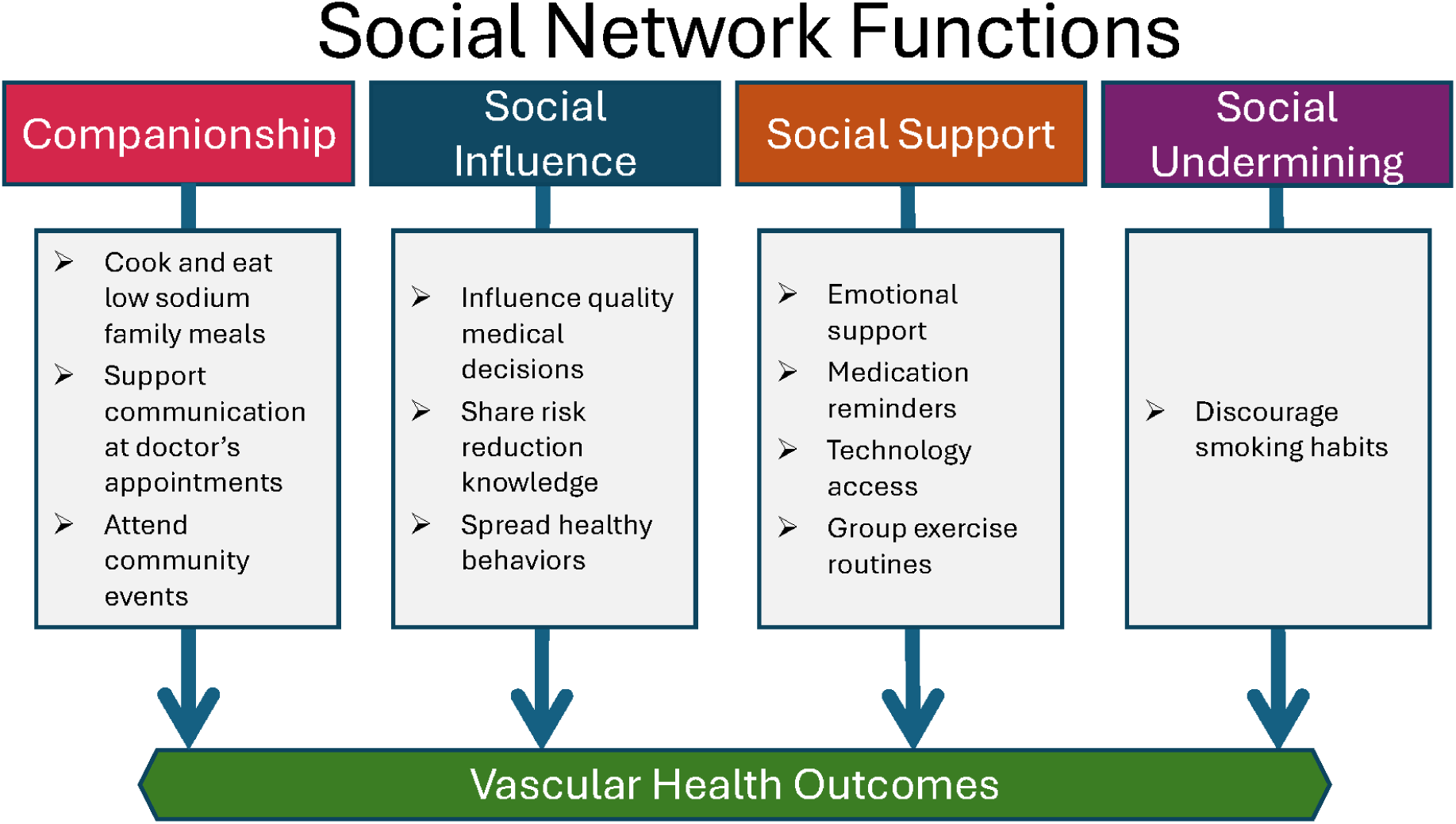
SERVE OC Conceptual Framework grounded in Social Network Theory. Adapted from Berkman et al.’s^47^ Social Network Theory framework, this model for SERVE OC outlines the hypothesized mechanisms linking social networks to vascular health outcomes. The schematic highlights core social network functions, including companionship, social influence, social support, and social undermining, along with specific operationalized examples for each domain within the SERVE OC intervention.

### 2.2 Study Setting

SERVE OC used a community engaged approach and included engagement of health systems, grassroot organizations, CHWs and dissemination plans which focused on ultimate implementation and transition to the community. We chose communities in Orange County with high rates of chronic disease.^50–53^ The study took place within the communities of Santa Ana, Anaheim, Garden Grove, and Westminster in Orange County, California. Orange County is home to Little Saigon, located in Westminster and Garden Grove, which has the largest Vietnamese community outside of Vietnam.^54^ Approximately one-third of Orange County’s population is Latino, with significant concentrations in cities like Santa and Anaheim.^55–58^ These local underserved communities face disparities within CVD due to historical and ongoing residential segregation of racial-ethnic minority groups, creating barriers to CVD prevention and management.^55,57^ As part of CBPR, we utilized CHWs and engage community stakeholders, local organizations, and other community members along all steps of the study process and intervention, which helped to identify barriers and tailor the intervention to Latino and Vietnamese communities in achieving health equity in CVD. We also specifically chose spaces out in the community to conduct this study. These spaces included El Centro Cultural De Mexico, a community-use space for predominantly Latino communities, Federally Qualified Health Centers (FQHCs) which are community centers built for underserved populations to access primary care and other health services, the Vietnamese-American Cancer Foundation which has community space tailored to local Vietnamese communities, and local parks and schools. Additionally, families were also offered help with transportation to-and-from appointments and childcare, if necessary. Both data collection and intervention appointments were conducted at community spaces or at-home based on participant preference, and included enrollment and follow-up.

### 2.3 Community Engagement in study design, implementation and dissemination

SERVE OC was grounded in a philosophy of community engagement with a focus on meaningful and sustainable partnerships. CBPR methodologies were used to adapt the study design including the family-based intervention to local Latino and Vietnamese communities. These strategies included engaging a Community Advisory Boad (CAB) comprised of CHWs, community partners, and other community stakeholders in focus groups and activities to integrate community engagement into every aspect of this RCT. Community engagement was applied to advance health equity and reduce health disparities by directly addressing SDOH, identifying community-level barriers and facilitators to CVH, and foster community-driven solutions. We worked with community partners to help us identify and hire CHWs, identify optimum recruitment strategies, and host and activate community events. Focus groups were conducted to help adapt the study design, inform study logistics, culturally tailor the intervention and identify barriers and opportunities in conducting a family-focused heart health intervention. Focus groups were conducted with CAB members, local health care providers, and Latino and Vietnamese community members during the planning and adaptation of the intervention, which continued throughout the first year of SERVE OC. The insight from focus groups helped to identify several barriers to CVH family interventions including access, feasibility, resources, and education.^50^ CAB meetings were held to discuss focus group findings and identify topics, strategies, and family-based activities to integrate into study procedures and the intervention. The research team then worked closely with CHWs to review intervention materials, incorporate and culturally tailored the information and activities, and establish protocols and a list of community resources for CHWs to better support families. Towards the end of the study the CAB worked with the research team on dissemination efforts including an event reporting preliminary findings back to the community and producing a short, documentary style videos on SERVE OC.

#### 2.3.1 Community Health Workers

In SERVE OC, CHWs were identified and hired from existing community organizations that we have long-standing relationship with. As in the DESERVE RCT, bilingual CHWs liaised between families, study staff, and community partners, and helped tailor intervention materials to ensure linguistic accuracy and cultural congruency.^15^ In SERVE OC, CHWs were fluent in multiple languages (English, Vietnamese, or Spanish), dependent upon the community they work with, which helped optimize recruitment, engagement, and retention. While CHWs had previous local organization training, as part of SERVE OC, CHWs underwent 2–3-hour trainings per day over the course of two to three days prior to the start of the trial, and refresher trainings as needed. CHW training content included review of intervention materials, motivational interviewing skills, data collection protocols, clinical trial and study procedures to aid with recruitment strategies, and the basics of CVH, including accurate BP measurements. CHWs, working alongside our research coordinators, conducted in-person family intervention sessions and assisted adult participants using the validated Withings BPM Connect device to enable home-based blood pressure readings as recommended by the U.S. Preventive Services Task Force (USPSTF).

### 2.4 Participant Population, Recruitment, and Sampling

This study was focused on testing a family intervention as a solution to addressing heart health in two large, underserved communities (Latino and Vietnamese populations) within Orange County, which have previously faced systemic barriers and bear high burden for CVH. Latino and Vietnamese populations represent a significant portion of the Orange County community.^55,56,58^ Recruitment took place from November 2022 up until April 2024, with a recruitment goal of 200 to 220 families. To reflect the population distribution of these underserved communities, we aimed to recruit 70% Latino and 30% Vietnamese families.^54,55,57–59^ We started with a cluster randomized sampling scheme to recruit Latino and Vietnamese families. We later adjusted our recruitment strategies by integrating convenience sampling and geographic cluster sampling to better reach Latino and Vietnamese families within the Santa Ana, Westminster, Garden Grove, and Anaheim area Recruitment efforts included targeted door-to-door outreach with a team consisting of study staff and CHWs. Recruitment strategies expanded to include holding information tables at local community events like local markets, health clinics, food distribution centers, parks, and FQHCs, which was successful in reaching potential participants to achieve our recruitment goal. To increase the number of Vietnamese participants, participants were recruited through referrals from community partners via telephone and email.

### 2.5 Participant Screening, Eligibility, and Consent

During recruitment, participants were screened for inclusion into the study using the criteria shown in Table 1. Operationally, families were defined as at-least two related individuals living within the same household and individuals are at least five years old. Informed consent was obtained from all participants before their enrollment within the study. For compensation, families received $50 per adult per family at each visit: baseline, 12 months, 24 months, and 36 months.

**Table 1.**
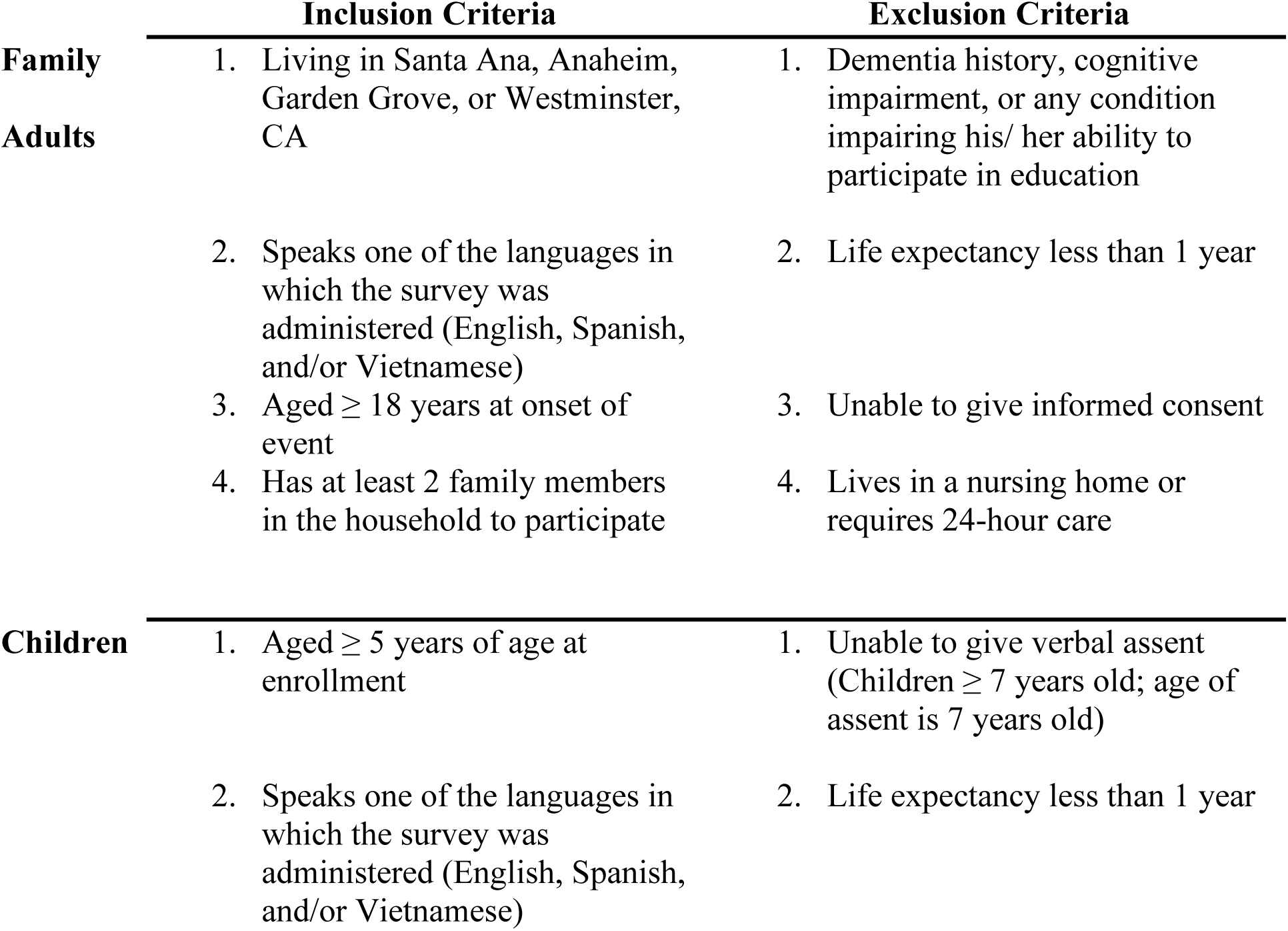
Inclusion & Exclusion Criteria for SERVE OC.

### 2.6 Primary Outcomes and Measures

The first primary outcome was the difference in proportion of “ideal” LE8 scores between the SERVE OC intervention and ISM treatment groups at 12, 24, and 36-month follow-ups.^46^ The second primary outcome was the SBP reduction difference (in mmHg) between adults in SERVE OC intervention and ISM groups at 12, 24, and 36-months. These outcomes are measured by AHA’s LE8 score, which is a holistic and standardized tool for assessing CVH by analyzing two domains: health behaviors and health factors.^46^ Within these 2 domains there are 8 individual factors which contribute to CVH: diet, physical activity, BMI (weight), nicotine exposure, sleep health, blood glucose, blood lipids, and BP.^46^ Each LE8 factor score ranges from 0 to 100 points, with higher scores reflecting a better state of CVH outcomes. Total LE8 score is calculated by adding all individual LE8 factor scores together and dividing by 8. “Ideal” LE8 was categorized as a score of ≥80.^60,61^ Measurements and calculation methods for each factor are described in greater detail in Supplemental Table 1, adapted from Lloyd-Jones et al.^46^

Other outcomes included social network data, cognitive assessment, and SDOH. Secondary outcomes included 1) the role of social networks at the family-level in modifying LE8 scores in the intervention versus ISM arms, 2) the impact of cognition in modifying LE8 scores in the intervention versus ISM arms and 3) the impact of SDOH (continuous and multiple SDOH (≥3 domains) on LE8 in both arms. For social networks, we measured the generations within the household, household size, family-type, and social support. Cognition was assessed using the Montreal Cognitive Assessment (MoCA), which is a brief, useful and validated cognitive screening measurement developed to screen for Mild Cognitive Impairment (MCI) and other cognitive disorders.^62,63^ SDOH was measured through five key domains using seven measurement indicators: economic stability (assessed through Medicaid enrollment), education (indicated by completion of 8th grade education), social connection (measured by confidence in receiving regular help from others), healthcare access (evaluated through regular healthcare source and insurance status), and geographic disadvantage (assessed using Area Deprivation Index and Social Vulnerability Index quartiles using census tracts).

### 2.7 Data Collection

Data was collected from individual family members at baseline and during 12-, 24-, and 36-month follow-ups by study staff and community health workers through staff-led interviews, bio-measurements, and self-reported surveys during in-person appointments. Dietary assessment was conducted through food frequency questionnaires including Dietary Approaches to Stop Hypertension (DASH) and Mediterranean Eating Patterns for Americans (MEPA), that evaluated adherence to nutritional guidelines.^64,65^ Physical activity scores were calculated based on self-reported weekly moderate to vigorous activity minutes, with scores assigned proportionally from 0 to 100. BMI scores were derived from measured height and weight using a standardized scale and stadiometer, with points assigned based on standard BMI categories. Nicotine exposure was evaluated on a scale considering current use status, quit history, and environmental exposure, with point deductions for household smoking exposure. Sleep health was scored based on self-reported nightly sleep duration, with optimal ranges (7-9 hours) receiving higher scores. Blood lipids were measured by levels of non-HDL cholesterol (mg/dL) collected through a fingerprick (capillary) sample. Blood glucose control was evaluated through HbA1c testing via fingerprick, and the scores reflected different levels of glycemic control. BP scoring incorporated both SBP and diastolic blood pressure (DBP) (mmHG), taking the average of two to three BP measurements using a validated BP monitor. At baseline, contact data, socio-demographics, biological measurement data, social network data, clinical data (medications, cognitive assessment, medical history), and behavioral history were collected through surveys. Cognition was assessed using MoCA, which screens for cognitive function by assessing 7 domains of cognitive function including orientation, attention, verbal memory, language, visuospatial function, and executive memory; it is a preferred tool for identifying executive dysfunction, and cognitive domains associated with cerebrovascular disease.^62,63,66^ As a proxy for mental health, depressive symptom severity was measured using the Center for Epidemiologic Studies-Depression (CES-D-10) scale, which is a validated and abbreviated 10-item measuring tool scored from 1-30.^67^ A full list of survey measures and timeline of measurement collection on LE8, socio-demographics, social networks, health history, access, and behavior, healthy literacy and SDOH, and psycho-social factors are further listed in Table 2. All gathered data was recorded on paper and/or electronically. Additional BP measurements were collected remotely through the validated Withings RBPM given to participants for at home readings, and data from their device transfers automatically to the Zot platform.^68^

**Table 2.** Timeline of Data Collection and Study Measurements.

| <u>Variable</u> | <u>Assessment Tool</u> | <u>Baseline</u> | <u>12</u><br><u>Months</u> | <u>24</u><br><u>Months</u> | <u>36</u><br><u>Months</u> |
| --- | --- | --- | --- | --- | --- |
| **LE8** |  |  |  |  |  |
| Diet | LE8<sup>46</sup> | X | X | X | X |
| Physical Activity |  | X | X | X | X |
| Nicotine Exposure |  | X | X | X | X |
| Sleep Health |  | X | X | X | X |
| BMI (weight & height measurements) |  | X | X | X | X |
| Blood Lipids (Cholesterol) |  | X | X | X | X |
| Blood Glucose (HbA1C) |  | X | X | X | X |
| BP (SBP, DBP) |  | X | X | X | X |
| **Social Networks, Support, & Demographics** |  |  |  |  |  |
| Socio-Demographics: Contact Info, Age, Sex, Gender, Race/Ethnicity, Income, Employment Status, Education, Country of Birth, Years in Community, Marital Status, Disability, Preferred Language, Preferred Language Match with PCP/ Doctor, Members of Household | Investigator-Adapted Socio-Demographic Questionnaire<sup>97-99</sup> | X |  |  |  |
| Social Networks | Investigator-Adapted Social Networks Alters Form<sup>100</sup> |  | X | X | X |
| Family Dynamics & Function | McMaster FAD – General Functioning Scale<sup>101</sup> |  | X | X | X |
| **Health History, Access, and Behavior** |  |  |  |  |  |
| Medical Insurance | Investigator-made Questionnaire | X | X | X | X |
| CVD/Medical History and Vascular Health Follow-Up Outcomes |  | X | X | X | X |
| COVID-19 History |  | X | X | X | X |
| Medication Inventory |  | X | X | X | X |
| Tobacco Use | CDC BRFFS Tobacco<sup>102</sup> |  | X |  | X |
| Alcohol Use | CDC BRFFS Alcohol<sup>102</sup> |  | X |  | X |
| Physical Activity | IPAQ<sup>103</sup> |  |  | X | X |
| Medication Adherence | MMAS-8<sup>104</sup> | X | X | X | X |
| **Health Literacy and Social Determinants of Health** |  |  |  |  |  |
| Health Literacy | BRIEF Health Literacy Screening Tool<sup>105</sup> |  |  | X |  |
| CVH Health Knowledge | Investigator-Adapted from the Stanford Five-City Project<sup>106</sup> |  | X | X |  |
| Discrimination | Major Experience of Discrimination (Abbreviated Version)<sup>107</sup> |  |  | X |  |
| Food Insecurity | Food Insecurity – PhenX Toolkit Protocol (PX 270301)<sup>108</sup> | X | X | X | X |
| **Psycho-Social Factors** |  |  |  |  |  |
| Cognition | MoCA<sup>62,63</sup> | X | X | X | X |
| Sleep | MOS<sup>109</sup> | X | X | X | X |
| Health Locus of Control | MHLC Health Locus of Control<sup>110</sup> |  | X | X | X |
| Depression | CESD-R-10<sup>67</sup> | X | X | X | X |
| Social Support (child) | CASSS (child only)<sup>111</sup> |  |  | X |  |
| Sedentary Activity/Screen Time (child only) | CHIS for Adolescents<sup>112</sup> |  | X |  |  |
| Perceived Stress | PSS-4<sup>113</sup> |  | X | X | X |

### 2.8 Study Enrollment and Randomization

After consent was obtained and participants were deemed eligible for inclusion, they are enrolled into the study. Enrolled families were each given a unique family ID number (code) for the study. If not all eligible family members joined during the initial enrollment, additional family members were allowed to be enrolled later during the study, joining their unique family ID number and the same treatment group as their family members for consistency and to avoid intervention contamination.

After enrollment, all families had their baseline measurements taken by study staff which included a brief interview, survey intake form, and bio-measurements. Adult participants were given a Withings RBPM (one per family), walked through device set-up, and given further instructions on how to accurately take their BP weekly. After baseline assessments are completed, families were randomly assigned to the intervention group (SERVE OC) or the usual care (ISM) group using stratified block randomization with block sizes of 4 and 6. For each language, the randomization was separate. Randomization lists were generated prior to beginning enrollment and replenished once to account for additional enrollment of families. Random block sizes of 4 or 6 ensure that the balance of families in intervention groups is never more than 3 families different.

### 2.9 Intervention Arm

#### 2.9.1 Family-Based Intervention - SERVE OC

Families randomized to the ISM group received standard of care, which consisted of language-appropriate LE8 information. Families randomized to the SERVE OC intervention group received standard of care plus a CHW-led family-based intervention focused on increasing vascular risk perception, family goal setting with lifestyle and activity planning, and skill building (medication adherence, provider-patient communication, nutritional literacy) to improve CVH through enhanced LE8. After the baseline appointment, CHWs connected with families to schedule three in-person family meetings within the first six months, followed by bi-weekly phone calls and/or messages from CHWs for the duration of the study. A study hotline was always available for families to communicate with CHWs outside of sessions. For the intervention CHWs were involved in: 1) Interviewing and recording of the family’s CVH risk factors through LE8 score and conversations with families on preventative measures, 2) Facilitating family conversation around an action plan which engaged the family network, 3) Providing support and advice to families to reinforce these plans via the SERVE OC app, and 4) Continued connection with families to motivate participation in ongoing community activities. The CHW model supported the family network through multiple avenues. Motivational Interviewing training was given to CHWs to facilitate conversations around barriers and challenges experienced by the family. CHWs tailored intervention sessions towards specific barriers that family networks might be facing and provided personal support. Families also received access to the SERVE OC app, which provides a platform for communication to their assigned CHWs, goal tracking, and culturally tailored resources on CVH like our SERVE OC Motivational Lifestyle Narrative videos. The intervention leveraged the family dynamic to motivate healthy changes in behavior through the structure of the social network, social support, and social diffusion.^47^ Education on LE8 and CVH provided to enrolled family members may facilitate the diffusion of health information within families, encouraging additional family members to participate in the study and adopt sustainable health behavior changes.

Intervention families were also invited to frequent community events, with at least one larger quarterly event held by study staff, community partners, community members, and CHWs at local venues to identify barriers and solutions to CVH while building community. These events were comprised of cooking classes, dance classes, art workshops, walking groups, community gardening, yoga, Zumba, soccer, and other active engagements. Families reported in-person for follow-ups and data collection at baseline, 12 months, 24 months, and 36 months. Detailed intervention components, delivery protocols, and implementation specifications are reported according to the Template for Intervention Description and Replication (TIDieR) checklist (Supplementary Table 2).^69^

### 2.10 Data Analysis

#### 2.10.1 Sample Size Estimation

For SERVE OC, the household was designated as the clustering unit. To ensure conservative power estimates, we assumed an intraclass correlation coefficient (ICC) of 0.5, such that study power would be greater if within-household correlations exceeded this value. Power analysis was conducted using several simulations with a fixed power of 80% and varied alpha levels of 0.05 or 0.05/3 to account for multiple testing across 12-, 24-, and 36-months assessments. We assumed an average household size of 5 and evaluated a range of overall prevalences for achieving 4 or more ideal LE8 factors at 36 months (0.2, 0.3, and 0.4) with a fixed sample size of 150 families (75 per arm). Based on our most likely scenario of a 30% prevalence in our control group, we estimated the study would have 80% power to detect an absolute between-group difference 17-19% difference (improvement) in prevalence of achieving 4 or more ideal LE8 factors at 36 months, depending on whether adjustment for multiple comparisons was applied.

For the secondary outcome of SBP, power calculations indicated that a sample of 375 subjects per arm would allow us to detect a minimum 9.6 mmHg reduction with 80% power (alpha = 0.05). These calculations assumed an ICC of 0.5, both within-subject and within-cluster correlations of 0.5, and standard deviations of 25 mmHg (between-subject) and 10 mmHg (within-subject). Given the conservative nature of these assumptions, our study is adequately powered to address our secondary aim of assessing changes in SBP, including less than a 10mmHg change. All calculations of sample size were performed using the clusterPower package in R.^70^

#### 2.10.2 Statistical Analysis

Randomization occurred at the household level, while the individual-level data served are the unit of analysis. SERVE OC’s primary analysis will account for within-individual and within-household correlations using mixed-effects logistic regression models which are suitable for hierarchical data.^71^ The first primary outcome, or the change in prevalence of four or more ideal LE8 factors over 12, 24, and 36 months, will be assessed using a time-by treatment interaction term to test differences in secular trends between the ISM and SERVE OC groups. Models will be adjusted for relevant covariates to minimize the influence of intra-cluster correlation. The second primary outcome of SBP will be analyzed using a linear mixed-effects model which will follow the same approach. These analyses will be completed when data collection closes.

#### 2.10.3 Baseline Analyses

We conducted analyses of the baseline data to describe our study population demographics, health characteristics, and LE8. Descriptive statistics were used to analyze race-ethnicity, age, education, employment status, preferred language, and disability status stratified across age groups and population, using means/ standard deviations or frequencies/ percentages. Additionally, descriptive statistics analyzed between-group differences in LE8 factors, depression, and MoCA scores stratified by age groups and intervention groups at the individual-level, and by language and intervention group at the family-level. Social networks were assessed through family-level descriptive statistics that analyzed family-type, people in the household, number of generations within the family, education, SDOH, food-security, and family-level LE8 scores. For MoCA analyses, scores were treated continuously, as well as categorically, using the recommended cutoffs and categories as described (0-18 Mild Alzheimer’s Disease (AD); 19-25 MCI; 26+ Cognitively Normal).^66^ The maximum score one can receive on the MoCA is 30 points – a sum of points for each MoCA domain. Additionally, one point was given to individuals who have twelve years or fewer of formal education (e.g., 12^th^ grade or less); however if a participant scores 30 points out of the total 30 points, but has 12 years or fewer formal education, an extra point is not added.^62,66^ A cutoff of 26 points was used to differentiate normal cognition from MCI, which has an average score of 22 and range of 19-25. Additionally, a cutoff score of 18 differentiates MCI from moderate cognitive impairment.^66^ At this cutoff score, the MoCA has 83% and 94% sensitivity to detect subjects with MCI and Mild AD, respectively.^63^ However, it is important to note that there is overlap in the scores for MCI and moderate cognitive impairment, since by definition, AD is determined by the presence of cognitive impairment in addition to loss of autonomy. As such, the MoCA team reports an average MoCA score for Mild AD to be 16, with a range of 11-21.^66^

We also conducted individual-level and family-level regression analyses to examine associations between SDOH, food security, and CVH measured by LE8 overall scores. At the individual level, we included adult participants identified as either Latino or Vietnamese. The study examined five key domains of SDOH, including economic stability, education, social connection, healthcare access, and geographic disadvantage. Each domain was categorized as either present or absent based on these indicators. To understand the cumulative impact of SDOH, we calculated the total number of SDOH barriers for each participant (ranging from 0 to 7 to reflect the collected variables) and categorized participants into two groups: those with <3 barriers, or ≥ 3 barriers. This allowed for examination of accumulated effects of social determinants across intervention and population groups at the individual and family-level. Food security was a family-level measure and was thus constant across adult household members for individual-level analysis. Linear regression models used continuous LE8 scores as the outcome variable, and logistic regression models modeled the odds of ideal CVH (LE8 ideal vs. not ideal). Exposures included count of SDOH barriers as a binary variable (≥3 vs. <3), and food security as a binary status (low security vs. high/marginal security). All individual-level models adjusted for age (mean-centered), sex at birth (reference: male), and population group (reference: Vietnamese). Family-level models used household-aggregated adult LE8 and SDOH measures as outcomes and exposures, respectively, and adjusted for household size and population group (reference: Vietnamese). Analyses were conducted in SAS software, Version 9.4 [Windows] and R Statistical Software 4.3.2.^72,73^

### 2.11 Data Management

Data were collected and managed through REDCap (Research Electronic Data Capture), a verified secure web-based platform for survey creation and data management, hosted at University of California, Irvine.^74,75^ Surveys were completed online through self-report or study staff administration. Routine data quality assurance procedures included data audits, recovery of missing data, correction of data entry errors and dual-review verification of measure scoring, to ensure accuracy and completeness. Once randomized, all households with adult participants 18 years and older were provided a Withings RBPM that allowed multiple users profiles. During set-up, participants were instructed on how to accurately perform home-based measurements and to take their BP at least once per week. Data from Withings RBPMs were collected and stored in ZotCare, an online research platform/database that connects to all Withings RBPMs and tracks weekly BP readings from families.^68^ ZotCare, available as Centralive, is a convenient HIPAA-compliant research platform for mobile and wearable health data.^76^ Lastly, point of care HbA1c and lipid panel devices were tested according to device instructions using multi-chemistry quality control solutions to ensure devices produced accurate readings.

## 3 Results

A total of 730 families were screened within these communities, 209 families completed enrollment and 521 did not meet eligibility and were excluded. An additional 19 families were excluded due to logistical factors including being unable to reach again, declined interest, missing an additional family member to qualify, and missed Withings RBPM set up. 190 families were randomized to either SERVE OC or ISM treatment groups. A Consolidated Standards of Reporting Trials (CONSORT) flow diagram (Figure 3) shows the flow of participants from recruitment to randomization.^77^ Of those families that were randomized, 141 were Latino families, 47 were Vietnamese families, and 2 were designated as “Other” populations. The ISM group was constituted of 70 Latino families, 25 Vietnamese families, and 1 “Other” family and the SERVE OC intervention group was comprised of 71 Latino families, 22 Vietnamese families, and 1 “Other” family. The ISM group had 243 individuals total, 187 adults and 56 minors whereas the SERVE OC intervention group had 243 individuals, 187 adults and 56 minors.

**Figure 3.**
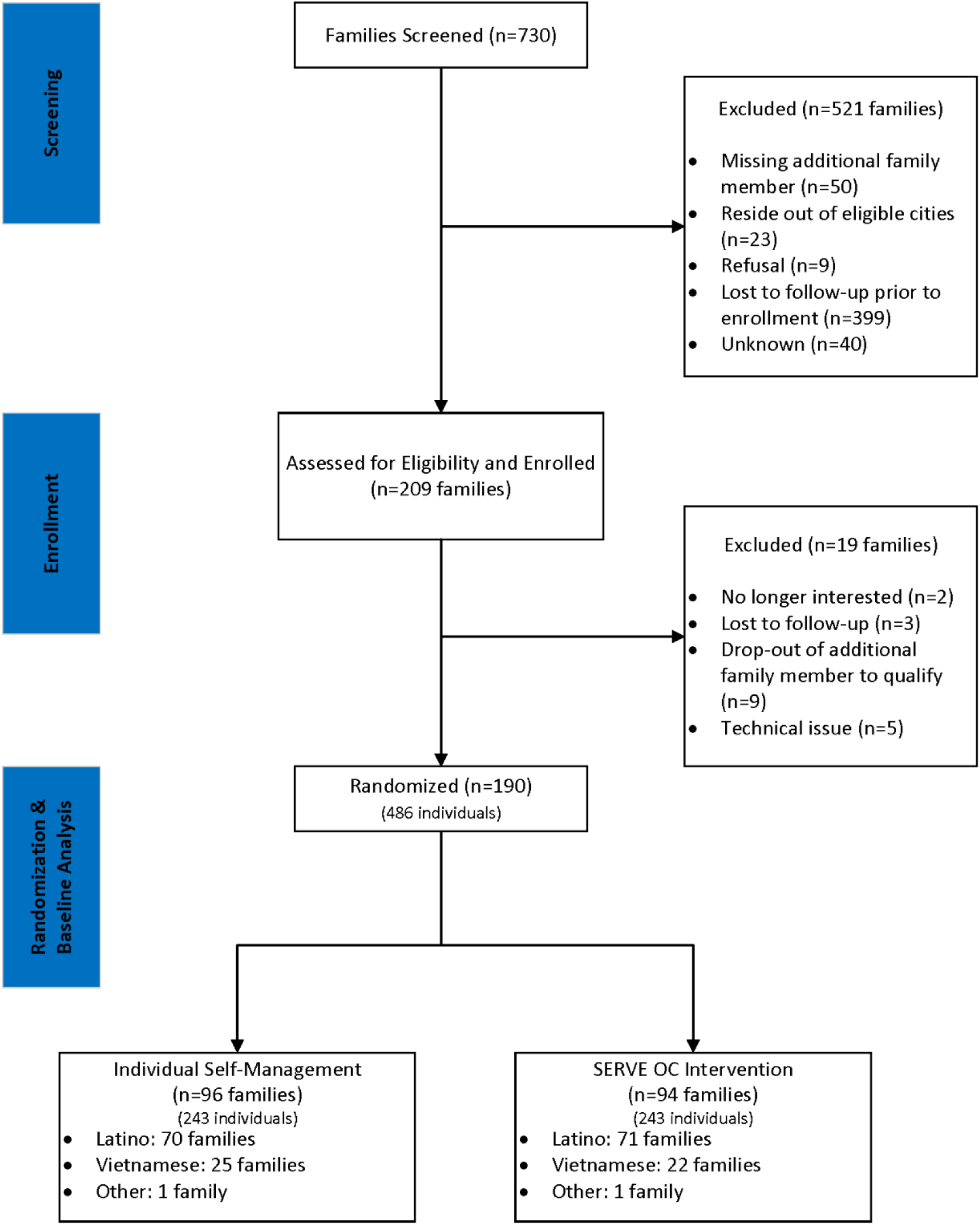
SERVE OC CONSORT Diagram for Participant Flow. Flow of participants through screening, enrollment, randomization, and baseline data analysis, modified from the Consolidated Standards of Reporting Trials (CONSORT) 2010 statement to reflect baseline allocation prior to intervention follow-up.^77^ Note: Follow-up and final primary outcome analysis stages are excluded as this report is restricted to methods and baseline data.

### 3.1.1 Baseline Findings

Characteristics for baseline demographics are shown in Table 3, stratified by age group and intervention group. Of all participants, the mean and median age for adult participants is 49.8 years (SD=15.7) and 50.0 years [18.0-87.0]. Over 76% of participants identified as Hispanic/Latino, whereas 22% identified as Asian, and about 2% as White. Spanish and Vietnamese were the preferred language of over 76% of participants. Almost half of adult participants had less than or equal to an 8^th^ grade level education (48%), and 70% were employed. Among all participants, 29% were born in the US and 64% born outside (7% missing).

**Table 3.**
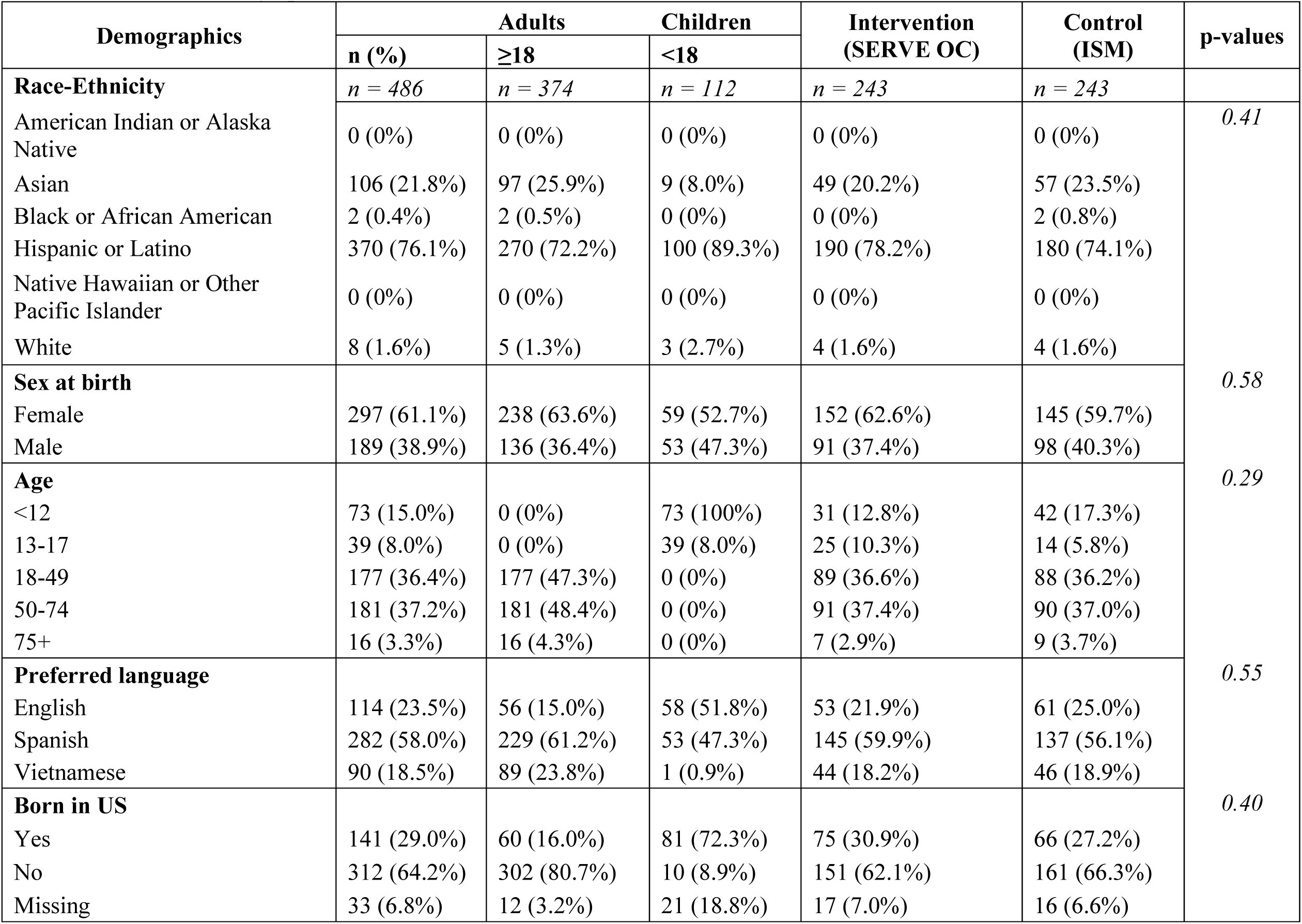
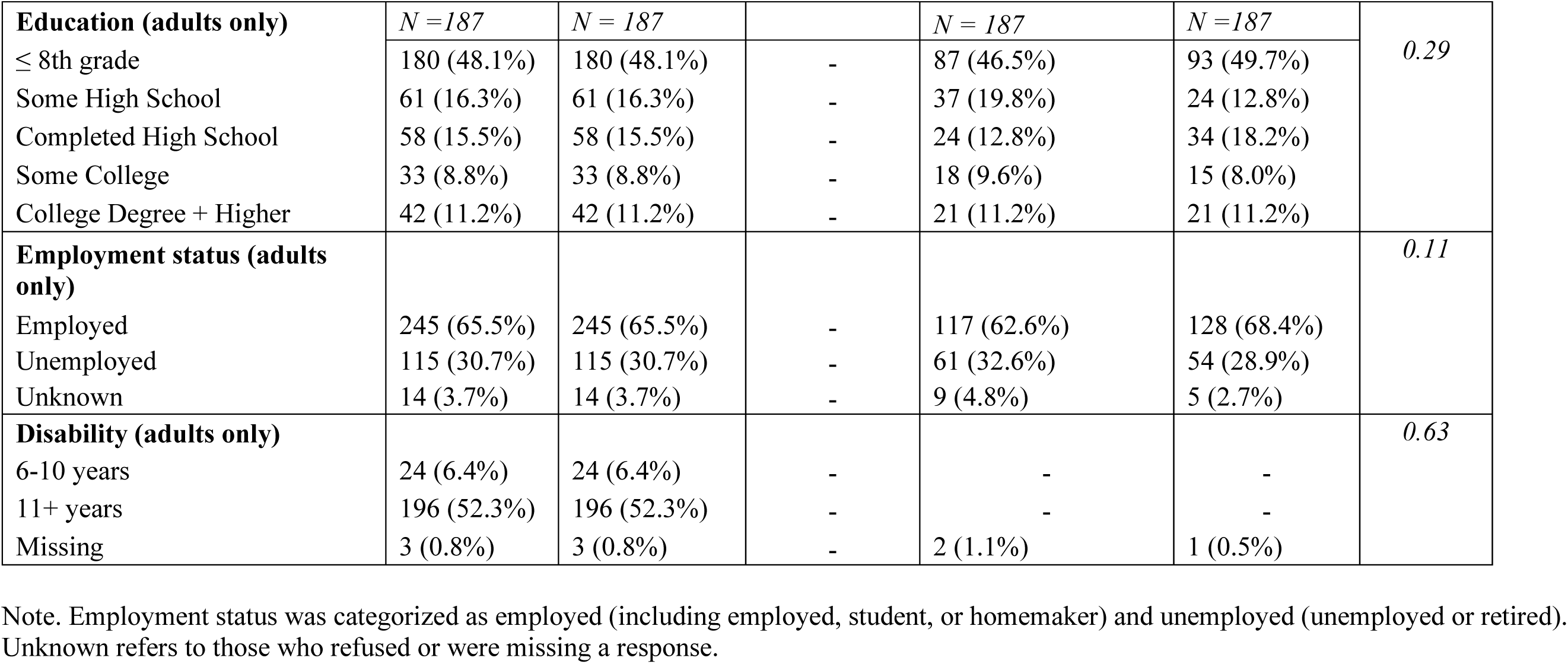
Baseline Demographics Results.

Baseline individual-level LE8, depression, and MoCA scores per each subgroup of age and treatment group are reported in Table 4. For over 83% of participants, LE8 scores were less than ideal health at baseline. Adults had an average total LE8 score of 66.61 (SD=11.96), compared to 76.52 (SD=10.15) in children. Average baseline LE8 total scores were comparable between adults in the intervention (67.57, SD=11.72) and adults in the control group (65.65, SD=12.16). Lowest average LE8 score for children was within the BMI domain (19.50, SD=12.34) whereas for adults, the lowest average LE8 score was physical activity (47.73, SD=42.15). The highest average LE8 score across all domains was in exposure to nicotine, where the vast majority were in the ideal range with averages above 90. Average depression scores for the CES-D were 8.04 (SD=5.53) in intervention adults and 7.98 (SD=5.35) in adults in the control group. Average MoCA scores among adults were 22.39 (SD=4.28), and were similar in intervention (22.78, SD=4.06) compared to control groups (21.98, SD=4.69).

**Table 4.**
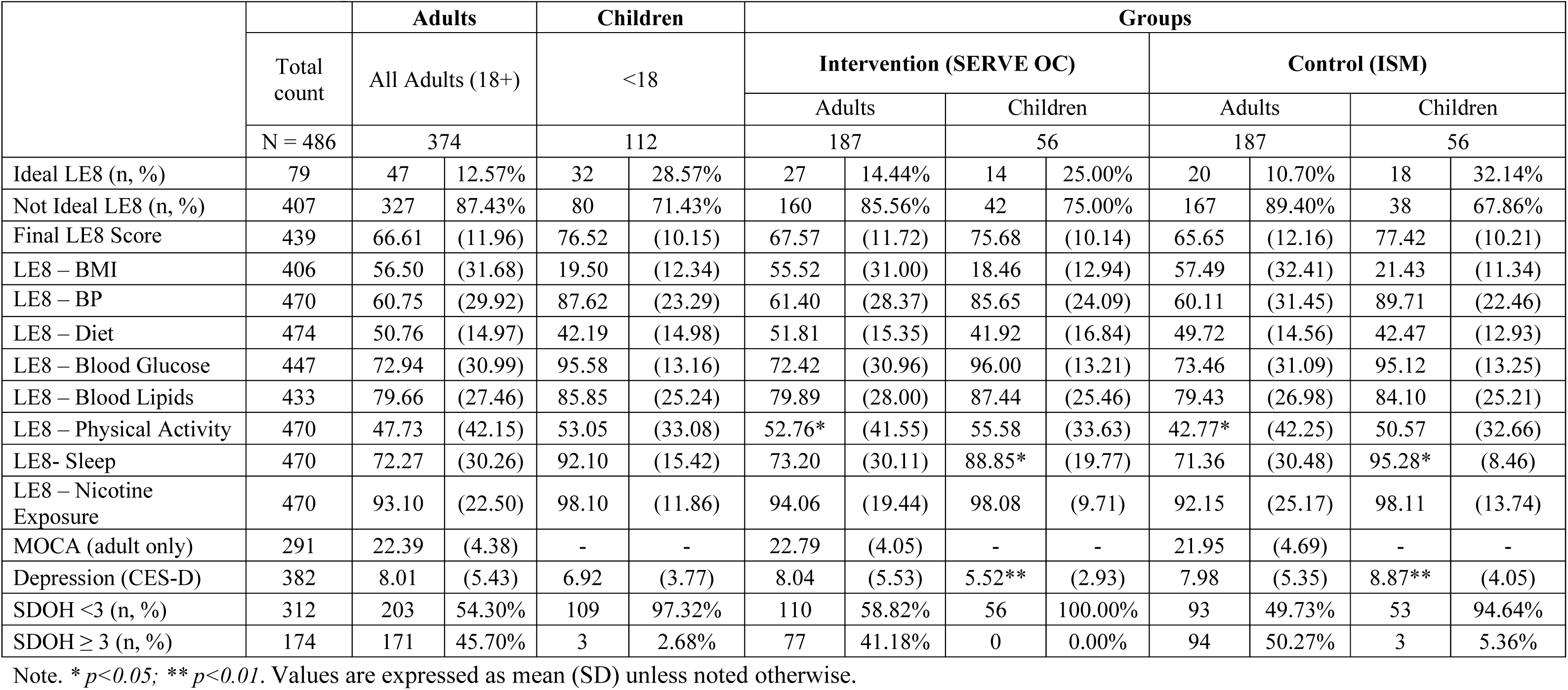
Individual-level LE8, Depression, SDOH, and MoCA Scores.

Regarding family-level LE8 scores at baseline, adults in the intervention families had a similar LE8 final score (67.38, SD=9.43) compared to adults in the control families (66.40, SD=10.08). Children in the intervention also had comparable baseline LE8 scores (75.65, SD=9.57) with children in the control group (76.73, SD=10.68) at the family-level.

At the individual level, a greater SDOH burden showed to be associated with worse CVH (Table 5). A higher SDOH barrier count was associated with lower LE8 scores (β = −1.93, 95% CI-4.40 to 0.53, p = 0.13) and lower odds of ideal CVH (OR = 0.55, 95% CI: 0.27 to 1.11, p = 0.09). Food insecurity was associated with lower LE8 scores (β = −2.92, p = 0.03) and lower odds of ideal LE8 (OR = 0.62, 95% CI: 0.32 to 1.22, p = 0.17). At the family level, no associations reached statistical significance, but the direction of the SDOH barrier associations was consistent with the individual-level pattern, with higher family SDOH barrier count associated with lower LE8 scores (β = −0.84) and lower odds of ideal CVH (OR = 0.57). Findings for family food security were directionally mixed: the continuous LE8 association was in the expected direction (β = −1.33, p = 0.47), whereas the ideal LE8 association pointed the opposite way (OR = 1.78, 95% CI = 0.42 to 7.58, p = 0.43) though higher food security score was consistently associated with higher odds of ideal LE8 (OR = 1.28, 95% CI: 0.95 to 1.73, p = 0.11).

**Table 5.** Logistic Regression Models of SDOH & Food Security.

| Level | Outcome | Exposure | Estimate Type | Estimate (95% CI) | p value |
| --- | --- | --- | --- | --- | --- |
| Individual | LE8 overall score | SDOH $\geq 3$ vs. SDOH $< 3$ | Beta | -1.93(-4.40,0.53) | 0.125 |
|  |  | Low vs. High/marginal food security |  | -2.92(-5.51,-0.32) | 0.029 |
| | LE8 Ideal | SDOH $\geq 3$ vs. SDOH $< 3$ | OR | 0.55(0.27,1.11) | 0.094 |
|  |  | Low vs. High/marginal food security |  | 0.62(0.32,1.22) | 0.168 |
| Family | LE8 overall score | SDOH $\geq 3$ vs. SDOH $< 3$ | Beta | -0.84(-4.18,2.50) | 0.622 |
|  |  | Low vs. High/marginal food security |  | -1.33(-4.91,2.24) | 0.466 |
| | LE8 Ideal | SDOH $\geq 3$ vs SDOH $< 3$ | OR | 0.57(0.15,2.20) | 0.414 |
|  |  | Low vs. High/marginal food security |  | 1.78(0.42,7.58) | 0.434 |

At the family-level (Table 6), among the 190 families enrolled in terms in the SERVE OC trial, 24% of families lived in a nuclear household structure, 65% lived in a multi-generational household and 10% lived in other or non-traditional structures. The mean household size was 4.34 individuals (SD=1.92) and 31% of households had children under 18 years of age. On average, 66% of all household members agreed to enrollment in SERVE OC. Mean family educational level among adults was approximately 10 years of education. On a scale of 0-7 domain measures, the mean number of SDOH burdens was 2.52 (SD=1.09) among the families. 83 families reported a greater SDOH burden ≥3, whereas 107 families reported <3 SDOH domains. Mean family food security score was 2.25 (SD=2.11), with 103 families reporting low food security and 54 families reporting high/marginal food security.

**Table 6.**
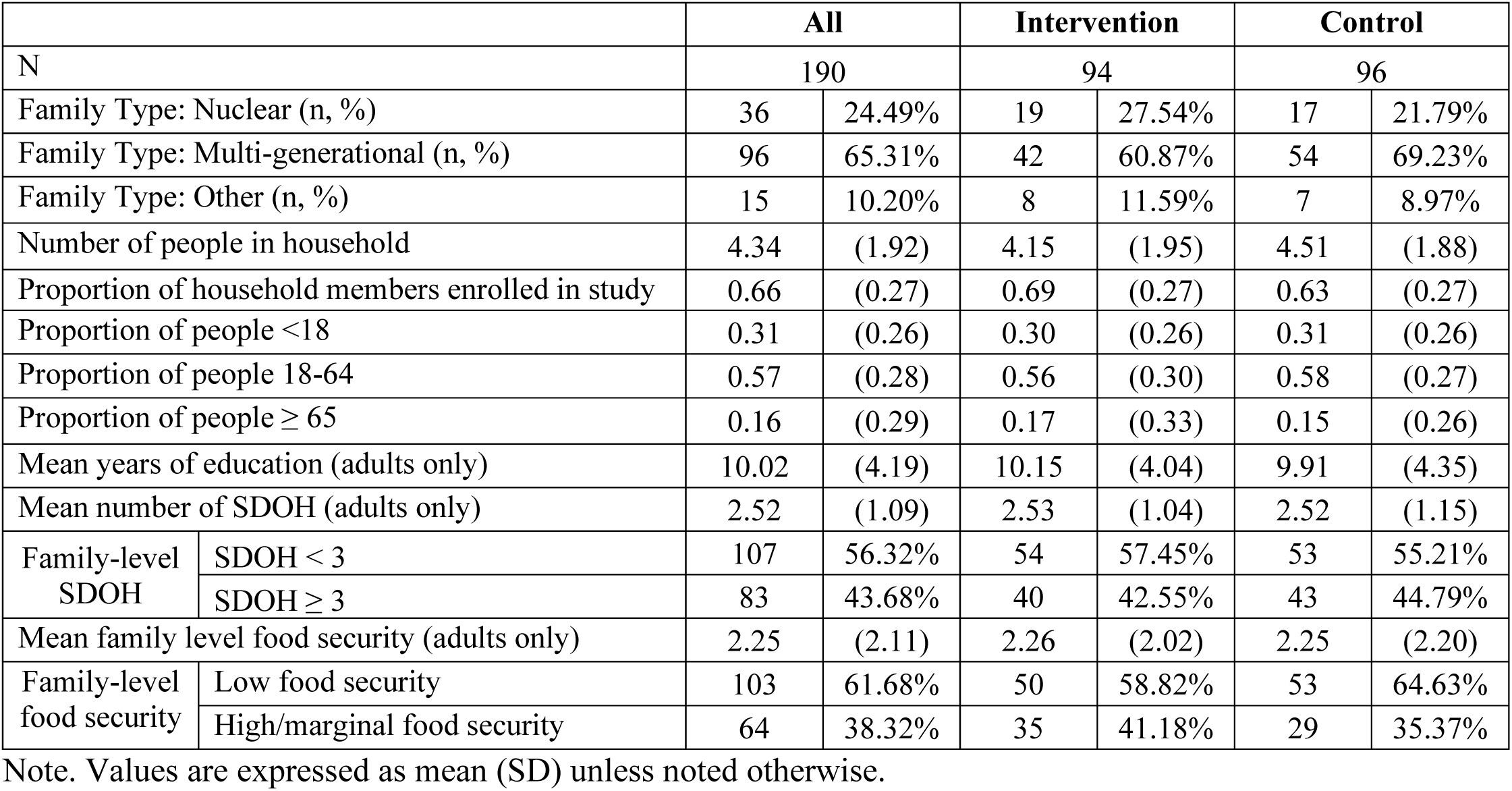
Family-Level Social Data.

|  |  | All |  | Intervention |  | Control |  |
| --- | --- | --- | --- | --- | --- | --- | --- |
| N |  | 190 |  | 94 |  | 96 |  |
| Family Type: Nuclear (n, %) |  | 36 | 24.49% | 19 | 27.54% | 17 | 21.79% |
| Family Type: Multi-generational (n, %) |  | 96 | 65.31% | 42 | 60.87% | 54 | 69.23% |
| Family Type: Other (n, %) |  | 15 | 10.20% | 8 | 11.59% | 7 | 8.97% |
| Number of people in household |  | 4.34 | (1.92) | 4.15 | (1.95) | 4.51 | (1.88) |
| Proportion of household members enrolled in study |  | 0.66 | (0.27) | 0.69 | (0.27) | 0.63 | (0.27) |
| Proportion of people $<18$ | | 0.31 | (0.26) | 0.30 | (0.26) | 0.31 | (0.26) |
| Proportion of people 18-64 |  | 0.57 | (0.28) | 0.56 | (0.30) | 0.58 | (0.27) |
| Proportion of people $\geq 65$ | | 0.16 | (0.29) | 0.17 | (0.33) | 0.15 | (0.26) |
| Mean years of education (adults only) |  | 10.02 | (4.19) | 10.15 | (4.04) | 9.91 | (4.35) |
| Mean number of SDOH (adults only) |  | 2.52 | (1.09) | 2.53 | (1.04) | 2.52 | (1.15) |
| Family-level SDOH | SDOH $< 3$ | 107 | 56.32% | 54 | 57.45% | 53 | 55.21% |
| | SDOH $\geq 3$ | 83 | 43.68% | 40 | 42.55% | 43 | 44.79% |
| Mean family level food security (adults only) |  | 2.25 | (2.11) | 2.26 | (2.02) | 2.25 | (2.20) |
| Family-level food security | Low food security | 103 | 61.68% | 50 | 58.82% | 53 | 64.63% |
|  | High/marginal food security | 64 | 38.32% | 35 | 41.18% | 29 | 35.37% |

Note. Values are expressed as mean (SD) unless noted otherwise.

## 4 Discussion

### SERVE OC Overview

SERVE OC was one of the first studies to examine the impact of a family-based intervention compared to enhanced self-management in elevating LE8 scores as a proxy for ideal CVH. While previous family-based interventions exist, many were limited by a dyadic paradigm centered on the patient and caregiver, positioning family members in a secondary supportive role.^78,79^ In contrast, SERVE OC diverted from the traditional individualistic or dyadic view of health behaviors. By involving the whole family, the goal of optimizing CVH is shared, with each member playing a distinct role in achieving better health outcomes for themselves and the family collective.^79^ SERVE OC advanced this concept by assessing the CVD risk of every participating family member using the LE8 framework. We hypothesize that identifying key lifestyle risk factors at the family level promotes both primary prevention for healthy members and secondary prevention for those with existing risk factors or documented vascular disease. The idea of testing a cardiovascular risk reduction intervention which simultaneously focuses on primary and secondary prevention using a family-based platform is novel.

This study also demonstrated the feasibility of enrolling entire families and utilizing CHWs to facilitate family-based initiatives, including lifestyle modification. Unlike other family studies, SERVE OC was designed to be accessible to underserved populations. This study has focused on Latino and Vietnamese communities of Orange County to successfully engage and recruit 190 families, including adults, children, and grandparents, with about two-thirds in multigenerational homes. These numbers demonstrate the feasibility of recruiting urban, underrepresented, and under-resourced families who face a disproportionate burden of CVD. While total participant counts fell short of our original target of 150 families (375 individuals) per arm due to smaller than anticipated average family sizes, our robust family-level recruitment highlights the success of this tailored outreach approach. Current literature focuses on a “dyad” dynamic, especially the patient-caregiver relationship around chronic illness, such as post-stroke and cardiovascular events.^78^ The literature reports significant burden is placed upon the caregiver, which can cause severe stress, physical health issues, and psychological strain.^80–83^ The dynamic of involving the entire family unit can help to alleviate caregiver burden, with the aim of improving overall functioning within the family.^79,84^ The family-based intervention model of SERVE OC aims to shift the paradigm from viewing the lifestyle behaviors as a burden to an opportunity to engage the entire family and solve the problem of poor CVH together.^79^ Our future data analyses will examine the effects of the intervention to improve CVH among family units.

Early and continued engagement with the community provided strategies which optimized enrollment and enhanced intervention development. For example, to enhance community involvement we rented space in a local, centralized community center, El Centro Cultural De Mexico, for recruitment and enrollment, in-person data collection, intervention sessions, follow-up appointments, and dissemination. These community-use buildings helped to break down barriers to inclusion by providing accessible, safe spaces to the community while minimizing time and resources on travel.^50^

This CHW-led family-focused intervention also highlighted the importance of integrating multiple community engaged strategies to enhance CVH education and promotion. Using CBPR principles, the intervention was adapted through focus groups and iterative review processes with community partners.^50^ Additionally, ideas from the community were implemented into the intervention’s community events and activities. Intervention events took place in the community with collaboration from local community organizations and included both SERVE OC-initiated and other existing programs, classes, and resources. By connecting participants to existing programs and classes, it provided sustainability for families to continue to engage with these resources after the SERVE OC study ends. Further, the partnership with the community helped initiate discussion around policy changes to promote better CVH through dissemination of findings to local government, community leaders, and the community itself. For example, if the community recommends creating a community garden to improve access to healthy foods, these findings will be presented to local leaders and organizations to develop an actionable plan for achieving this goal. While prior studies have incorporated elements of community engagement, including community-based activities and CABs, SERVE OC expanded upon these efforts by employing a multi-faceted approach. This work underscores the critical role of community involvement in in study design and implementation, as well as the importance of leveraging community strengths and resources to ensure long-term sustainability.

Similar to our prior experience in DESERVE, we employed CHWs to liaise between families, study staff, and community partners.^15^ Integrating CHWs into the study team provided us with a better sense of recruitment and retention feasibility, technological barriers, gaps in knowledge and education around CVH, and accessibility to resources amongst participating SERVE OC families. Our CHW’s gathered data around these challenges which allowed our study team to adapt to individual family’s needs in real time. Additionally, our CHWs played an important role in tailoring the intervention to specific Latino and Vietnamese cultures, thus making the intervention more culturally relevant. For example, CHWs worked with study staff in providing tailored, healthy recipe ideas that are culturally familiar and align with CVH dietary recommendations for our intervention materials and cooking classes. With a pre-established avenue into the community, CHWs are effective in engaging families and building trust which enabled the study’s success in recruiting and retaining multi-generational families from underserved communities. The relationships they built with the families in the study were crucial and demonstrate the importance of community engaged methods and culturally and linguistically congruent strategies.

Another important aspect of SERVE OC is the integration of technology within the intervention to reduce the digital divide and address health disparities faced by many underserved communities.^85^ This study focused on utilizing the SERVE OC app and RBPMs to translate data in real-time for CHWs, and leveraging the family dynamic to teach participants on technological literacy to improve health outcomes. RBPMs have been shown as a useful tool to help close the digital divide by providing participants with affordable, at-home devices to help keep track of their health.^85^ In addition, the use of RBPMs advanced digital literacy while aligning with USPSTF guidelines for home-based measurement to manage hypertension. However, SERVE OC is one of the first community-based studies to integrate family behaviors with health technology, in essence utilizing a reverse mentorship model in which younger participants help older generations navigate digital tools to directly advance health equity. Through the adoption of RBPMs and the family support structure, we aim to see significant longitudinal improvements in ideal LE8 scores and CVH.

Results from our baseline data collection demonstrate that we have yet to optimize CVH despite current prevention strategies, available treatments, and technological innovation like RBPMs and wearable devices. In our community-based cohort, only 13% of adults and 29% of children have ideal LE8 scores, with behavioral factors such as physical activity, diet, and sleep emerging as the largest contributors to poor CVH. Consistent with existing literature, our results further document the disproportionate burden of CVD, stroke, hypertension, and diabetes within underserved communities.^7,86,87^ Expanding upon these findings, SERVE OC focus groups revealed that financial constraints and limited access to healthy food options are primary barriers to maintaining ideal health.^50^ Our results support that food insecurity and lack of proper nutrition in these communities are not solely due to individual choice, but deeply rooted structural unavailability of affordable, nutritious food.^50,88^ Overall, the persistence of these modifiable risk factors despite the availability of pharmacological and technological innovation suggests that current prevention and treatment strategies are failing to adequately reach and support underserved populations.^86,89^ Consequently, this study focuses its efforts on optimizing ideal CVH through a family network-based intervention, leveraging social support to identify and address barriers specific to these underserved communities.

Additionally, our baseline results show patterns that are consistent with the literature on risk factors for CVH including cognition, SDOH, social isolation, and education. For instance, average MoCA scores among adult participants were low, which align with current research indicating that underserved communities face higher burdens of cognitive impairment.^90–92^ Moreover, participants experiencing a higher burden of SDOH, specifically total exposure to more than 3 adverse SDOH (social isolation/connection, lower educational attainment, economic stability, health care access, and geographic disadvantage/neighborhood/built environment) resulted in poorer LE8 scores. There is robust body of literature demonstrating that cumulative social disadvantage is associated with increased cardiovascular risk and poorer risk factor control.^93^ More specifically, results demonstrate that social isolation was associated with lower baseline LE8 component scores, including blood glucose, blood lipids, and sleep health. These findings also align with prior research linking social isolation to increased cardiovascular risk.^94^ Lastly, lower education attainment was associated with poorer LE8 scores, especially in BMI (weight management) and physical activity. When considered alongside our sample’s demographic profile, which reflects generally lower educational attainment, these findings provide important context for understanding the needs of the community we serve and highlight opportunities for targeted intervention. We have seen a similar pattern in our previous work, for instance, in DESERVE we found that family/friend networks with higher educational attainment were associated with greater reductions in SBP after 12 months of follow-up.^15^ Collectively, these results emphasize the continued need for tailored CVH education as well as the potential value of leveraging family friend networks to improve risk factor management and overall cardiovascular health.

Contrasting to the literature, participants showed minimal depressive symptoms as measured by the CES-D, with an average adult depression score of 8. Existing literature typically documents increased psychological distress in under-resourced populations.^95^ Therefore, additional analyses will be conducted to continue to examine depression overtime in our cohort.

Ultimately, our baseline findings reinforce the current cardiovascular landscape, demonstrating that despite the emergence of novel treatments, advanced technology, and an increased emphasis on prevention, CVH remains suboptimal in underserved communities. SERVE OC will assess whether a family-based intervention can serve as the necessary bridge to reduce persistent disparities in CVH. This approach represents a critical paradigm shift, moving beyond individual-level care to prioritize prevention at the family and community levels. Our results underscore the need for re-designing and implementing innovative prevention and treatment programs that can significantly enhance CVH for both adults and children, fostering a sustainable culture of healthy lifestyle behaviors within the home.

## 5 Limitations

Although our recruitment and baseline results highlight the feasibility of a family-based intervention, we did encounter challenges. Our inclusion criteria was focused on families that live in the same household, and a significant number of prospective participants had a large family-and-friend network that lived outside of the primary household. This study may have missed key individuals within these networks who are collectively invested in each other’s health successes. Thus, future research should consider expanding enrollment to include multi-household, family-and-friend networks to fully capture the influence of social connectivity. Due to the inclusion criteria, average eligible family units were smaller than projected, resulting in a final yield below our target enrollment of 150 families (n = 375 individuals) per arm. To account for smaller family sizes within the household, we recruited and randomized more families than expected to ensure a sample size that was adequately powered. Other issues to recruitment and enrollment included transportation, family schedules, staffing in preferred languages (Spanish and Vietnamese), and building trust within the community. These challenges added time and costs associated with recruitment and enrollment, and although they will not affect the results, they influenced our ability to meet our targeted enrollment timelines. Regarding data collection, the DASH survey did not accurately account for cultural dietary patterns, resulting in a positively skewed distribution of categorical responses for four dietary components (i.e., red meat, fish, butter, and sweets), caused by a scaling error in the determination of serving size cut points. To address this issue, sensitivity analyses were conducted and we chose to exclude these four components, instead focusing on the remaining six dietary components measured. Study staff collected both the actual number of servings reported by participants alongside the LE8 dietary categorical responses. Additionally, we added the 16-item MEPA questionnaire to supplement dietary intake data and mitigate any missing dietary information. Lastly, to mitigate time burden of participation, participants were initially sent home with a paper take-home survey and collected at later CHW visits. The survey consisted of measures on health literacy, CVH knowledge, major experience of discrimination, health locus of control, social networks (adult and children), tobacco, alcohol, and physical activity. However, this resulted in a low-response rate and was changed to in-person surveys during subsequent follow-ups to reduce the opportunity for missing data.

## 6 Conclusions

This study demonstrates the feasibility of enrolling entire families and utilizing CHWs to facilitate family-based initiatives, including lifestyle modification. Unlike other family studies, SERVE OC was designed to be accessible to underserved populations. Results from this clinical trial will help contribute to and support the evidence base for the use of culturally tailored, family-based interventions in reducing risk for CVD and improving CVH outcomes. Specifically, we seek to test whether a community-based approach to primary and secondary prevention of CVD, improves ideal LE8 scores in both Latino and Vietnamese intervention families. This multi-level clinical trial will directly utilize the results and feedback from the participating communities to inform community stakeholders, local government officials, and local organizations on solutions to barriers and obstacles faced by the local community, thereby initiating change at the policy level. To ensure these findings are translated into real-world practice, we have integrated the Reach, Effectiveness, Adoption, Implementation, and Maintenance (RE-AIM) framework to evaluate the readiness for implementation and the potential for long term scalability.^96^ SERVE OC results will be disseminated back into the community to help promote the adoption of researched-based strategies for CVD prevention. By fostering community and utilizing CHWs through the SERVE OC intervention, we hope to create sustainable avenues for connection and abundant educational resources for the local community that continue beyond the clinical trial.

## Data Availability

The data, analytic methods, and study materials that support the findings of this study are available from the corresponding author upon reasonable request.

## 7#Acknowledgements

We would like to dedicate this manuscript in loving memory of the late Dr. Bruce Albala, may his legacy carry on. We would like to acknowledge the NIMHD (Grant number P50MD017366) for their support of SERVE OC and the UC END Disparities P50 grant. We would like to thank the SERVE OC team. We would like to acknowledge Darnisha Draughter (SERVE OC team), Pilar Lara de Cortez and Guadalupe Capistran (Radiate Consulting), Guillermo Alvarez (Latino Health Access), Becky Nguyen and Dung Hua (Vital Care Access Foundation/Vietnamese American Cancer Foundation), Julia Bautista (El Sol Academy), Dr. Jose Mayorga and Leeanne Funada (UCI Federally Qualified Health Center) for their contributions to the SERVE OC study. We would also like to acknowledge our SERVE OC community health workers Maria Haddad, Margarita Ochoa, Hilda Martinez, Socorro Juarez, Quynh Do, and Jaimee Doan. Finally, we would like to acknowledge all our participants without whom none of this would have been accomplished.

## 8 Funding

This research was supported by the National Institute of Minority Health and Health Disparities, as a part of the National Institutes of Health funded P50 UCLA-UCI Center for Advancing Solutions to Cardiometabolic Health (CASCADE) (Grant No. P50MD017366).

## 9 Disclosures

The authors have no conflicts of interest to disclose.

## Non-Standard Abbreviations and Acronyms

AD: Alzheimer’s Disease
AHA: American Heart Association
BP: Blood Pressure
CAB: Community Advisory Board
CBPR: Community-based Participatory Research
CHW: Community Health Worker
CVD: Cardiovascular Disease
CVH: Cardiovascular Health
DASH: Dietary Approaches to Stop Hypertension
DBP: Diastolic Blood Pressure
DESERVE: Discharge Educational Strategies for Reduction of Vascular Events
FQHC: Federally Qualified Health Center
ISM: Individual Self-Management
LE8: Life’s Essential 8
MCI: Mild Cognitive Impairment
MEPA: Mediterranean Eating Pattern for Americans
MI: Myocardial Infarction
RBPM: Remote Blood Pressure Monitor
RCT: Randomized Controlled Trial
RE-AIM: Reach, Effectiveness, Adoption, Implementation, and Maintenance
SBP: Systolic Blood Pressure
SDOH: Social Determinants of Health
SEM: Socioecological Model
SES: Socio-economic Status
SERVE OC: Skills-Based Educational Strategies for Reduction of Vascular Events in Orange County
SNT: Social Networks Theory
SWIFT: Stroke Warning Information and Faster Treatment
USPSTF: U.S. Preventive Services Task Force

## 13 Supplementary material

Tables S1-S2

**Supplemental Table 1.** LE8 Factor Description, Method of Measurement, and Calculation Technique

**Supplemental Table 2.** Template for Intervention Description and Replication (TIDieR) checklist – SERVE OC

**References: #**46,64,65,69,103,114,115

## Notes

### Competing Interest Statement

The authors have declared no competing interest.

### Clinical Trial

NCT05641519

### Author Declarations

Ethical approval for this study was granted by the University of California, Irvine Institutional Review Board.

